# Role of pollution and weather indicators in the COVID-19 outbreak: A brief study on Delhi, India

**DOI:** 10.1101/2021.01.04.21249249

**Authors:** Kuldeep Singh, Aryan Agarwal

## Abstract

The present study examines the impact of environment pollution indicators and weather indicators on the COVID-19 outbreak in the capital city of India. In this study, we hypothesize that certain weather conditions with an atmosphere having high content of air pollutants, might impact the transmission of COVID-19, in addition to the direct human to human diffusion. The Kendall and Spearman rank correlation tests were chosen as an empirical methodology to conduct the statistical analysis. In this regard, we compiled a daily dataset of COVID-19 cases (Confirmed, Recovered, Deceased), Weather indicators (Temperature and relative humidity) and pollution indicators (PM 2.5, PM 10, NO2, CO, and SO2) in Delhi state of India. The effects of each parameter within three time frames of same day, 7 days ago, and 14 days ago are evaluated. This study reveal a significant correlation between the transmission of COVID-19 outbreaks and the atmospheric pollutants with a combination of specific climatic conditions. The findings of this research will help the policymakers to identify risky geographic areas and enforce timely preventive measures.

## 1. Introduction

Coronavirus disease 2019 (COVID-19) is an infectious disease which initially detected in Wuhan, China, has now spread all over the world, and if not well dealt, it could even lead to the worldwide economic crisis. This virus exhibits high human-to-human transmissibility that is the reason it has spread all across the world in a very short span of time. The World Health Organization (WHO) reported 4,789,205 COVID-19 confirmed cases and 318,789 deaths worldwide until May 20, 2020 [1]. The deadly virus has affected more than 210 countries had been affected where United States (US) alone contributed approximately one-fourth of the total cases. Other major countries having a significant impact are Russia, Spain, Italy, Iran, France, Germany, UK, Turkey, China, and many more. The situation in India has started worsening day by day. The first COVID-19 case in India was reported on January 30, 2020 and as on May 20, 2020 India has 106,750 COVID-19 confirmed cases, including 3303 deaths [1]. As per WHO, India has not entered into the state of the community transmission, still it has clusters of cases.

India had enforced mandatory nationwide lockdown and enforced stricter rules to combat the COVID-19 pandemic since March 25, 2020 and further extended it till May 31, 2020. The lockdown might have helped in combating the transmission rate of the COVID-19 pandemic. Figure 1 shows the overall picture of confirmed cases of COVID-19 across the states of India.

**Fig. 1.**
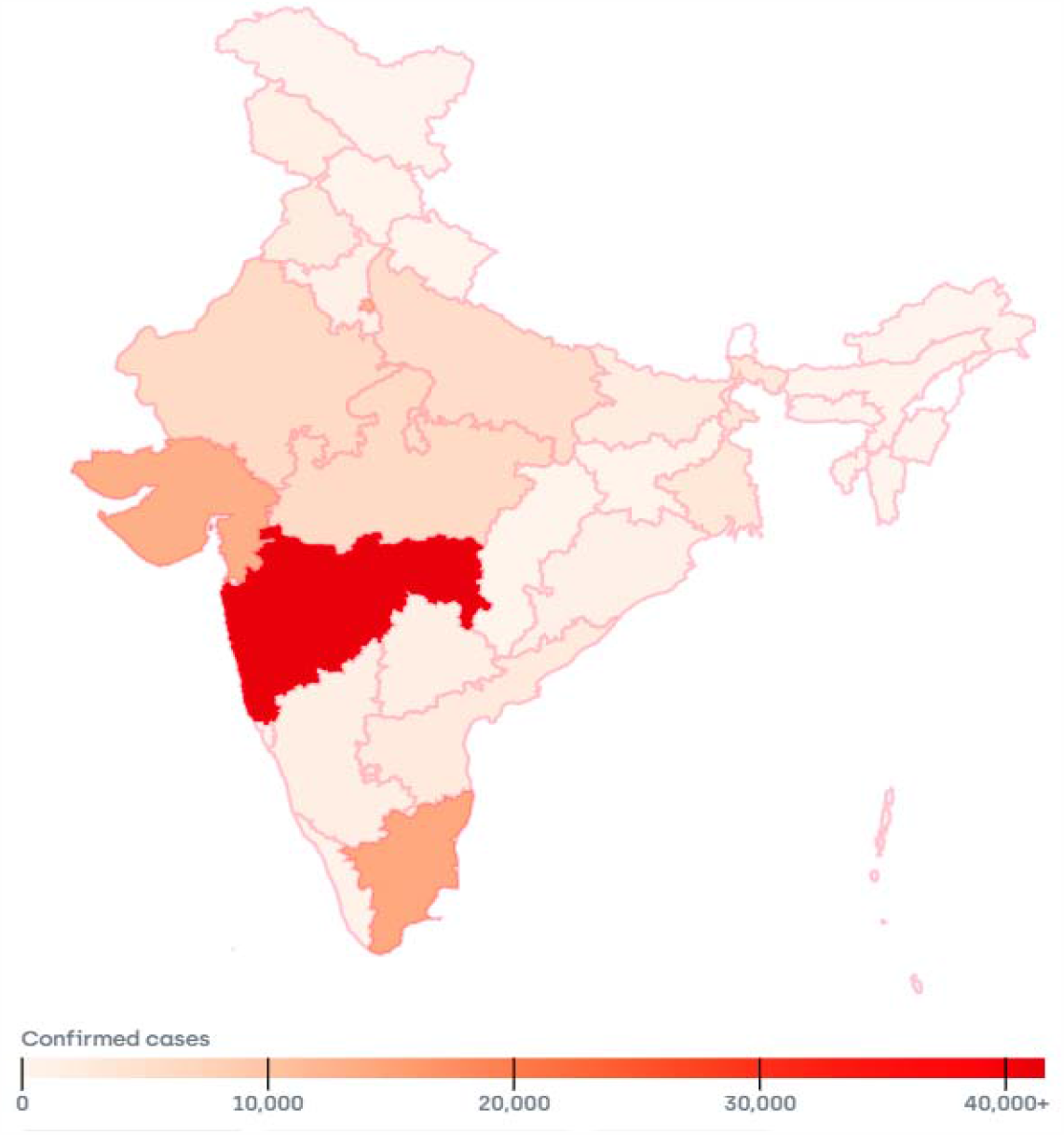
Outbreak of COVID-19 in India (Source: https://www.covid19india.org/)

Many geographical, social, political, and environmental factors might influence the impact of this deadly virus. Delhi is one of the most affected states in the India, currently has fourth highest infected number of patients. As of May 20, 2020, Ministry of Health and Family Welfare, Government of India has confirmed more than 10,500 cases, including>166 deaths in Delhi state. Delhi has experienced a drastic reduction in air pollution up to 50% [2] due to the implementation of extended mandatory lockdown to prevent COVID-19 transmission. Since, Delhi is considered among the most polluted cities of the world [3], it is worth analysing the role of environment pollution indicators on COVID-19 outbreak.

The link between exposure to air pollutants such as PM2.5 and PM10, Sulfur dioxide, nitrogen dioxide, carbon monoxide and ozone and increased risk for respiratory virus infections as well as susceptibility to respiratory virus infections is well established [4] [5]. There is significant evidence that air pollution is associated with influenza [6], pneumonia and acute lower respiratory infections [7], and severe acute respiratory syndrome [8]. Although much more about COVID-19 remains to be learned, some studies have tried to examine the air quality and impact of environmental pollutants’ association with COVID-19. Table 1 summarizes the findings of these studies conducted in various countries worldwide.

**Table 1:** Recent studies on the impact of pollution indicators on COVID-19 spread

| Ref. | Country | Pollution Indicators | Statistical measure | Outcome of Study | Duration of Study |
| --- | --- | --- | --- | --- | --- |
| [9] | Italy | PM<sub>10</sub> and PM<sub>2.5</sub> |  | SARS-CoV-2 could find suitable transporters in air pollutant particles | Feb,2020 |
| [10] | Italy | Air Pollution |  | the high level of pollution in Northern Italy should be considered an additional co-factor of the high level of lethality | Feb-Mar 2020 |
| [11] | China | PM<sub>2.5</sub>, PM<sub>10</sub>, CO, NO<sub>2</sub> and O<sub>3</sub> | Generative Additive Model | Positive associations of PM<sub>2.5</sub>, PM<sub>10</sub>, CO, NO<sub>2</sub> and O<sub>3</sub> and Negative association of SO<sub>2</sub> with COVID-19 confirmed cases | Jan 23 to Feb 29, 2020 |
| [12] | Italy, Spain, France and Germany | NO2 |  | long-term exposure to this pollutant may be one of the most important contributors to fatality caused by the COVID-19 virus | Feb-Mar 2020 |
| [13] | Italy | PM10 |  | The role of airborne PM for the virus diffusion is not evident | Feb 10 to Mar 27, 2020 |
| [14] | United States of America | PM2.5 | Negative binomial mixed model | We found that an increase of only 1 $\mu\text{g}/\text{m}^3$ in PM2.5 is associated with an 8% increase in the COVID-19 death rate | Jan 1 to Apr 22, 2020 |
| [15] | China | PM10 and PM2.5 |  | Higher concentration of PM10 and PM2.5 has a positive correlation with deaths caused by COVID-19 |  |
| [16] | Italy | NO2, O3, PM2.5 and PM10 | Pearson Correlation | Chronic atmospheric pollution may favour coronavirus spreading | Jan 1 to Apr 27, 2020 |
| [17] | United States of America | PM 2.5, PM 10, NO2, CO, Pb and SO2 | Kendall and Spearman Correlation Coefficient | Environmental pollutants such as PM10, PM2.5, SO2, NO2, and CO have a significant correlation with the COVID-19 epidemic in California | Mar 4 to Apr 24, 2020 |

**Table 2:**
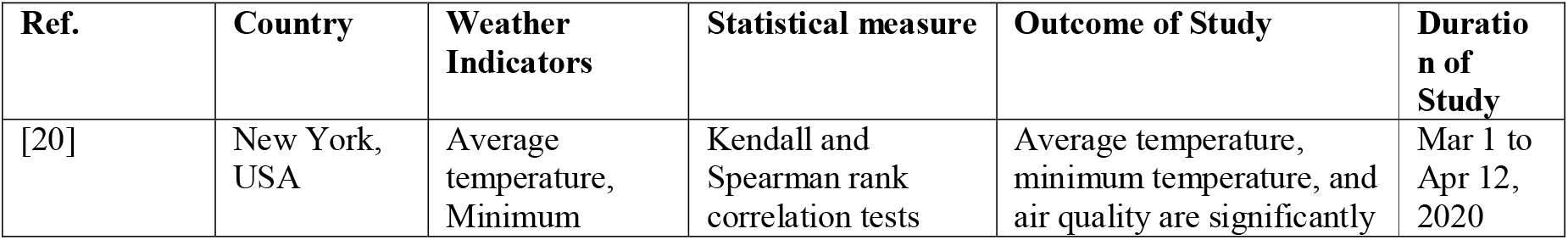
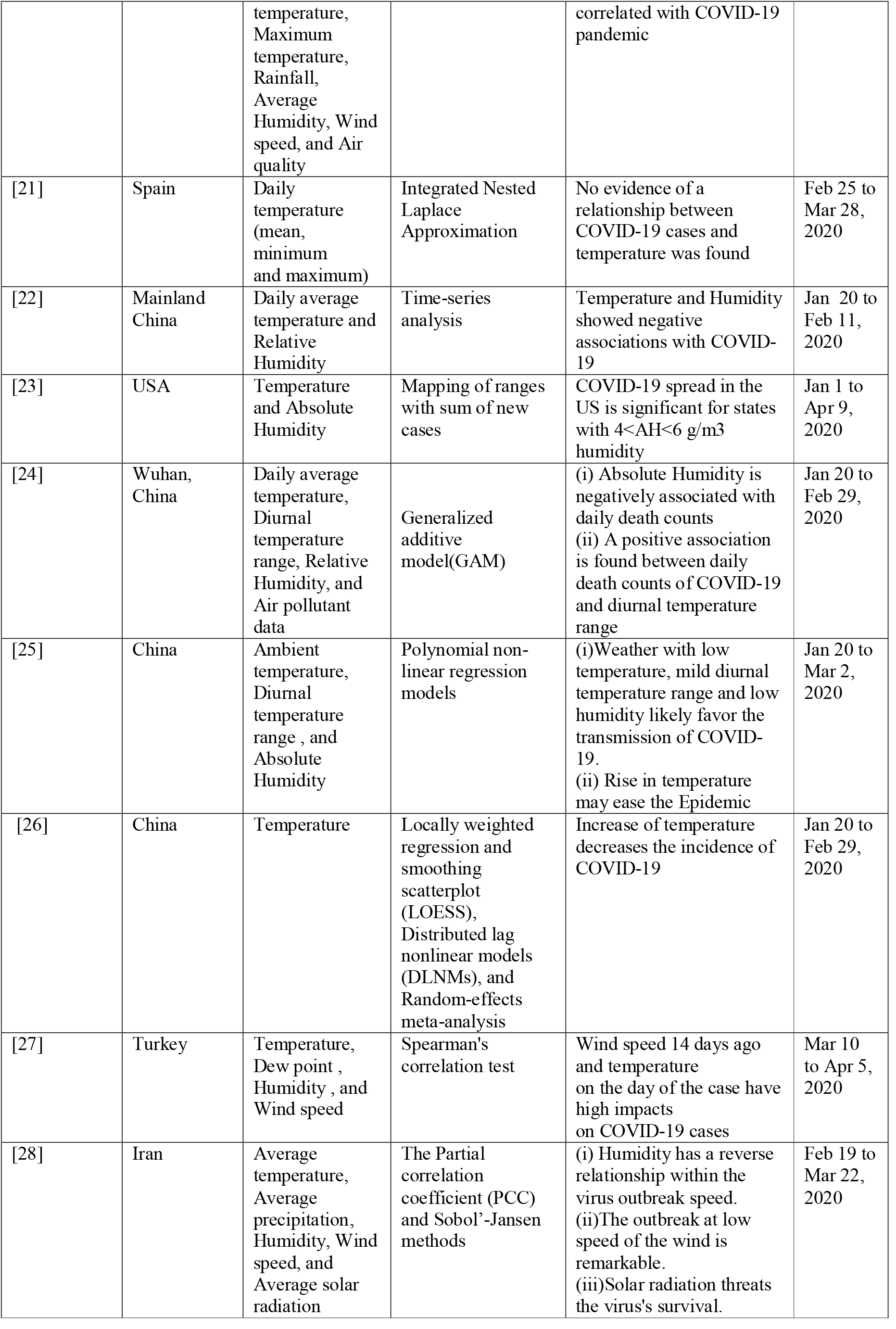
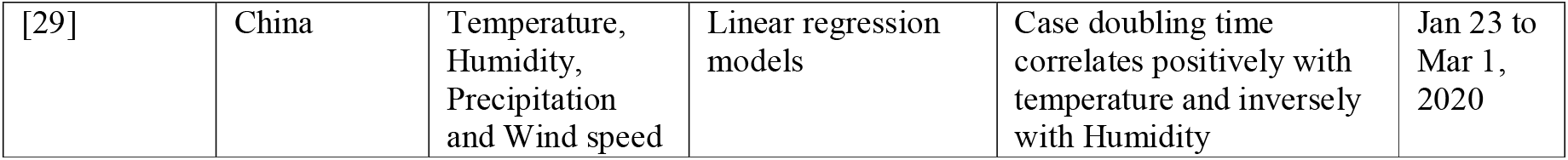
Recent studies on the impact of weather condition on COVID-19 Spread

Earlier studies have also suggested that the meteorological parameters might be active factors in the transmission of viruses and disease emergence. Yuan et al. investigated that the peak spread of SARS occurred in a particular range of Temperature, Relative Humidity, and Wind Velocity [18]. Variations of absolute Humidity correlate with the onset and seasonal cycle of influenza viral in the US. [19]. Recently, various studies have been conducted to analyze the impact of weather conditions on the spread and effect of COVID-19. We have summarized the outcome of these studies conducted worldwide in the Table2.

The independent effect of air pollution as well as weather indicators on the transmission of COVID-19 has not been studied systemically in the Indian context. Therefore, it is reasonable to speculate that environmental and meteorological factors might affect the spread of COVID-19 in India as well. In this paper, we attempt to answer a very important research question that need prompt responses: “Is there a potential association between the transmission of the coronavirus and the levels of air pollutants as well as weather indicators?” To the best of our knowledge, this is the first study to explore the effects of air quality and weather conditions on COVID-19 outbreak in India.

The primary weather indicators i.e. temperature (°C), and Humidity (%) and pollution indicators such as PM 10, PM 2.5, Sulfur dioxide (SO2), Carbon monoxide (CO), and Nitrogen dioxide (NO2) was considered as independent variables for finding the correlation with affected cases of COVID-19. The majority of the related recent studies have considered the indicators of the same day, which could present a false picture in terms of the correlation with COVID-19 cases. Since the incubation period of the COVID-19 virus varies from 1 day to 14 days, we have evaluated the impact of pollution and weather indicators for three time frames, namely on the day, 7 days ago, and 14 days ago.

## 2. Materials and Method

### 2.1 COVID-19 Data

The data of COVID-19 cases of Delhi state of India was retrieved from a publicly available repository and accessible through this link: https://www.covid19india.org/. This is a volunteer-driven, crowd-sourced database being collected and homogenized from multiple official and private web sources. In the dataset three parameters of COVID-19 cases are collected i.e. daily confirmed, daily recovered and daily deceased cases from Apr 1 2020 to May 19 2020 as highlighted in Figure 2.

**Fig. 2.**
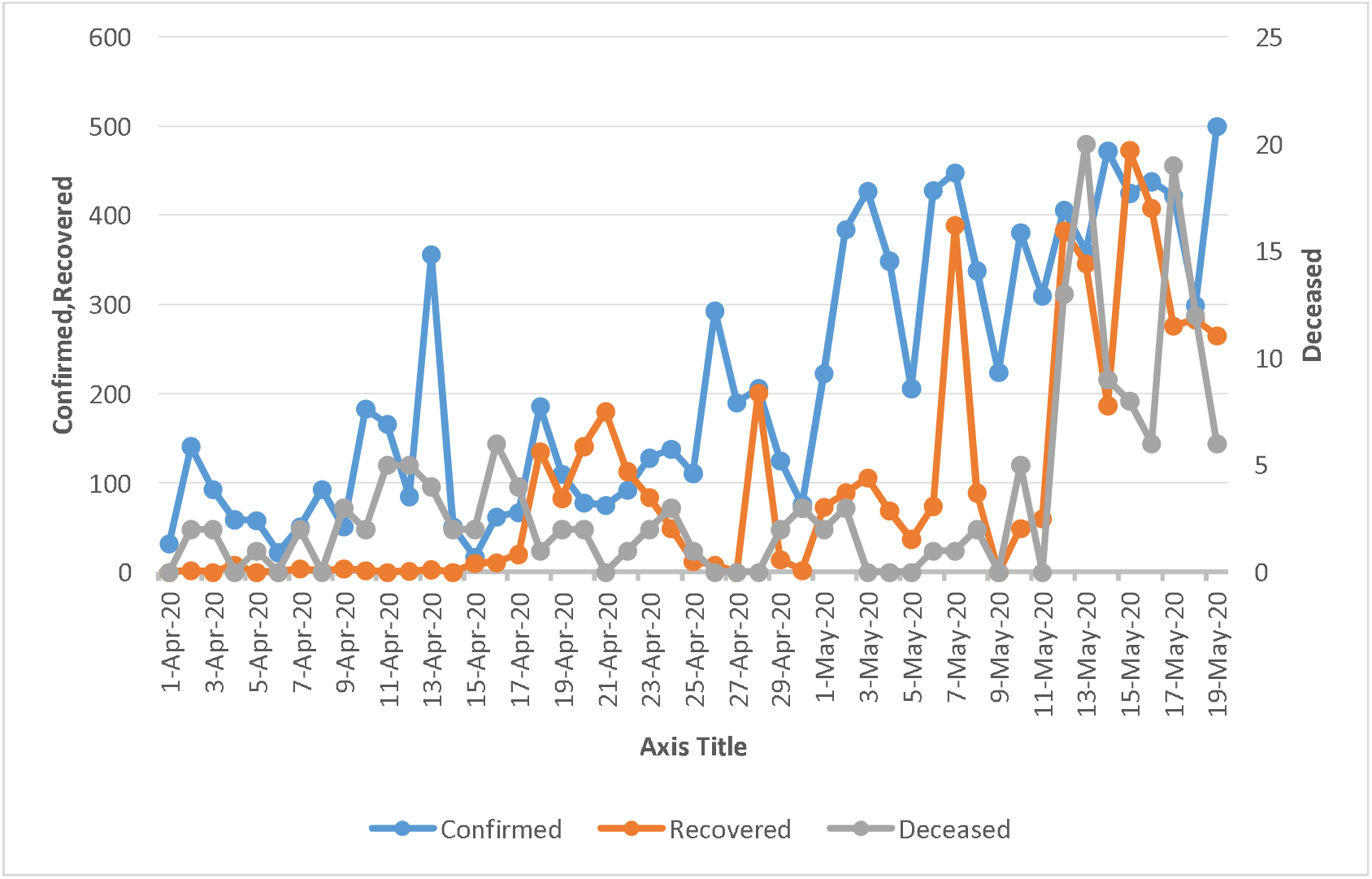
COVID-19 case statistics in Delhi

### 2.2 Pollution Data

The dataset of environmental pollution indicators such as PM 10, PM 2.5, Sulfur dioxide (SO2), Carbon monoxide (CO), and Nitrogen dioxide (NO2) was taken from OpenAQ Platform (https://openaq.org), which includes historical archives of pollution indicators observations from local government air quality agencies i.e. Central Pollution Control Board (CPCB). The duration of data collection was Mar 15 2020 to May 19 2020 to cater the time frame of 14 days old data. Figure 3 depicts the pollution profile in Delhi during the period of this study.

**Fig. 3.**
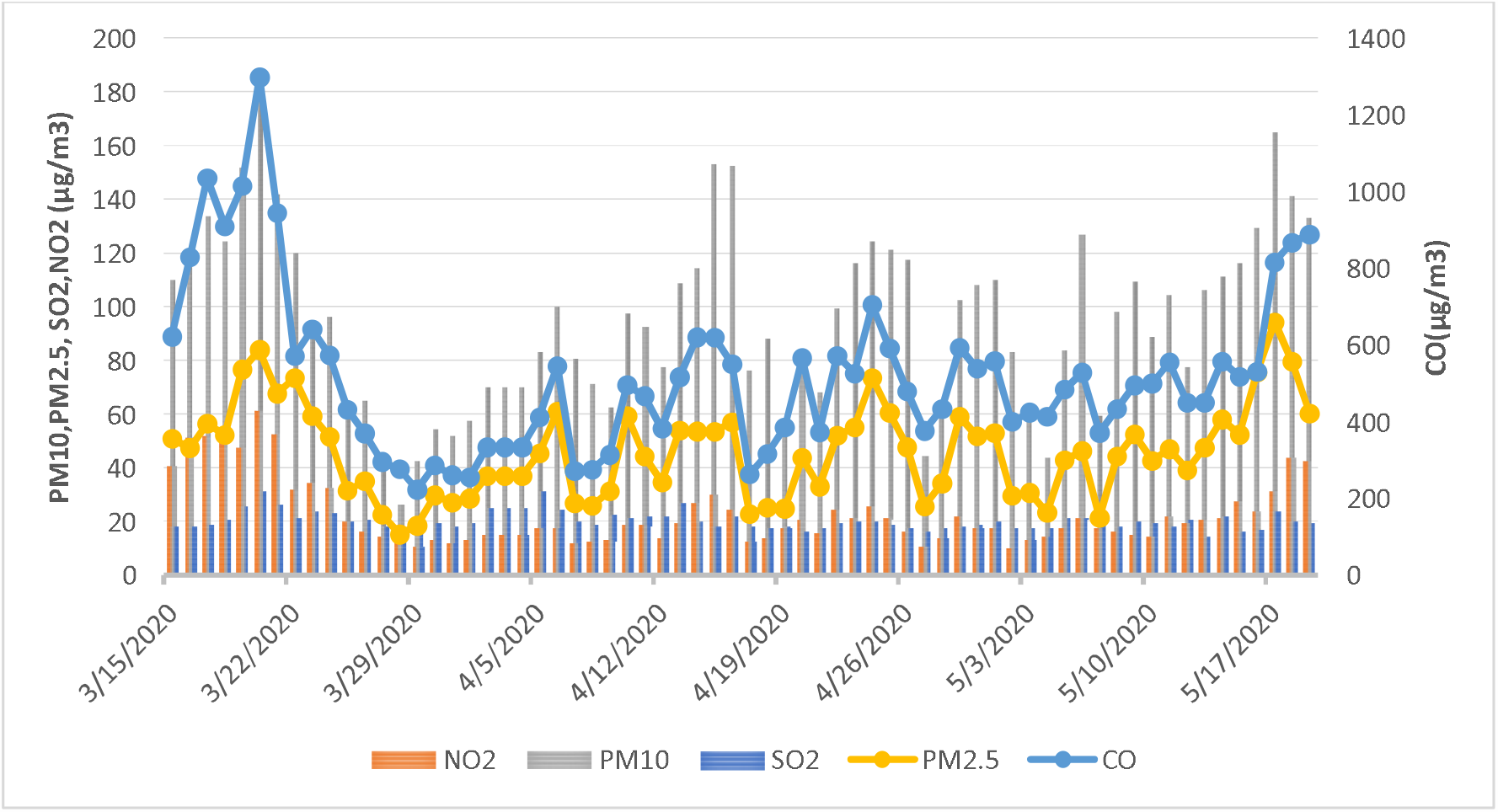
Pollution indicators statistics in Delhi

### 2.3 Weather Data

The meteorological data used in this study including Temperature (Max, Average), Humidity (Max, Average) were retrieved from Weather Underground (https://www.wunderground.com/) from Mar 15 2020 to May 19 2020.

### 2.3 Statistical analysis

As the data collected are not normally distributed; therefore Kendall [30] and Spearman rank correlation [31] tests are considered in this study to investigate the association of pollution and weather indicators with COVID-19 cases. Both of these methods are accepted measures of non-parametric rank correlations. We hypothesize that certain weather conditions with an atmosphere having high content of air pollutants, might favour the transmission of COVID-19, in addition to the direct human to human diffusion. A null hypothesis corresponding to each indicator is formulated that there is no association between individual indicators and the three case statistics considered in this study.

## 3. Results and Discussion

The significant findings of this study show that pollution and weather indicators are significantly correlated with COVID-19 daily confirmed, recovered and deaths in Delhi. Table 3 depicts the empirical estimations of environmental pollutants’ correlation with COVID-19. The study highlights that the concentration of Particulate Matter indicators PM10 and PM2.5 in two time frames i.e. same day and 7 days ago have a positive correlation with deaths caused by COVID-19. These findings are in line with the study conducted by Yao et al. [15] in China. PM10 and PM 2.5, 7 days ago are also significant for daily confirmed cases and the recovered cases. The impact of CO and NO2 same day and 7 days ago on daily recovered cases can be easily observed. SO2 values same day and 14 days ago has also shown impact on daily recovered cases and the correlation is negative similar to the findings of Zhu et al. [11] for China. Overall, the pollution indicators 14 days ago have negligible influence on the COVID-19 outbreak. Similarly, all the pollution indicators on same do not have any significant relation with daily confirmed cases. The results also indicate that the most reasonable timespan to study the impact of pollution indicators is 7 days ago.

**Table 3:**
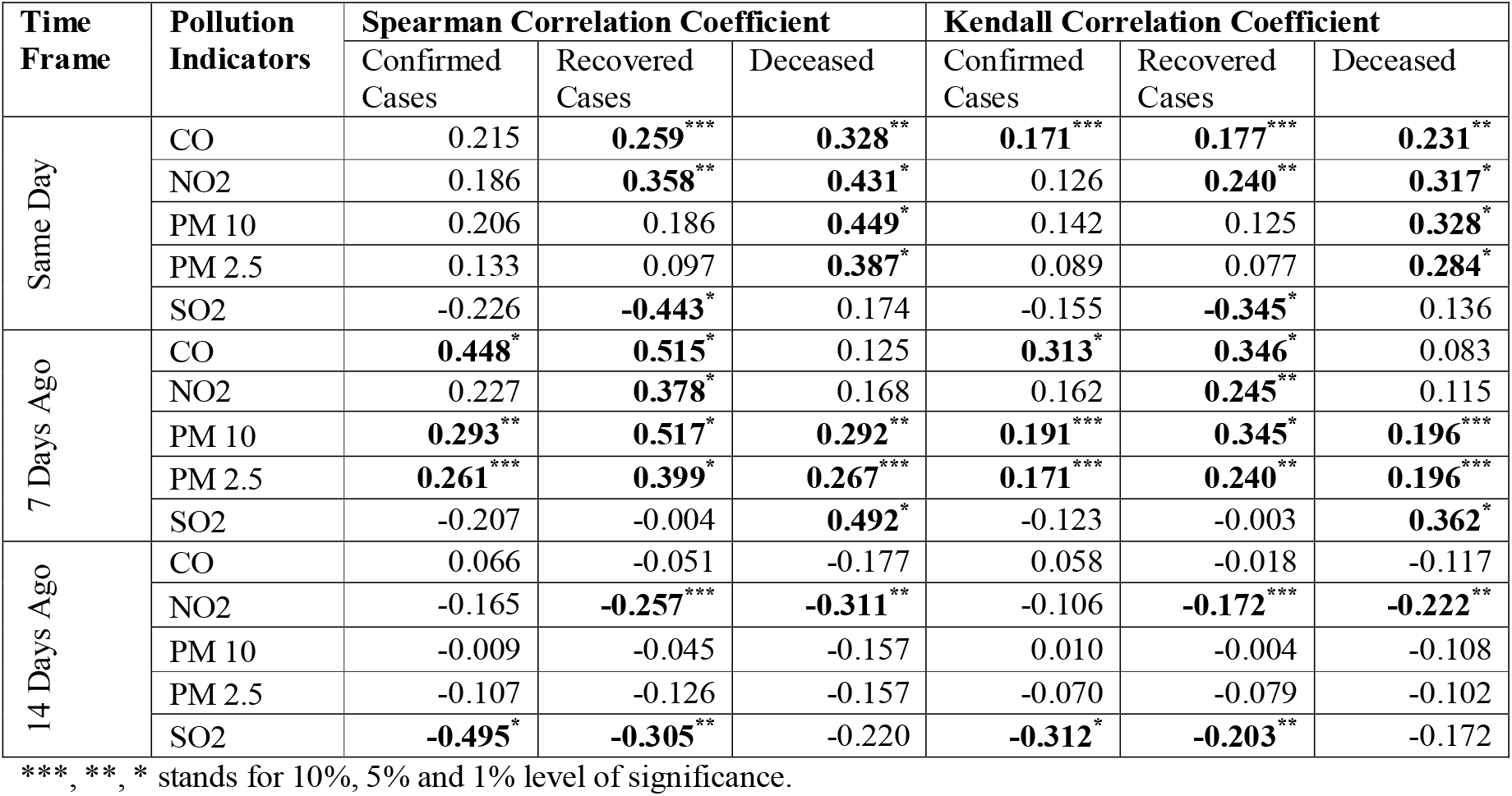
Correlation coefficients between pollution indicators and COVID-19 cases

| Time Frame | Pollution Indicators | Spearman Correlation Coefficient |  |  | Kendall Correlation Coefficient |  |  |
| --- | --- | --- | --- | --- | --- | --- | --- |
|  |  | Confirmed Cases | Recovered Cases | Deceased | Confirmed Cases | Recovered Cases | Deceased |
| Same Day | CO | 0.215 | **0.259\*\*\*** | **0.328\*\*** | **0.171\*\*\*** | **0.177\*\*\*** | **0.231\*\*** |
|  | NO2 | 0.186 | **0.358\*\*** | **0.431\*** | 0.126 | **0.240\*\*** | **0.317\*** |
|  | PM 10 | 0.206 | 0.186 | **0.449\*** | 0.142 | 0.125 | **0.328\*** |
|  | PM 2.5 | 0.133 | 0.097 | **0.387\*** | 0.089 | 0.077 | **0.284\*** |
|  | SO2 | -0.226 | **-0.443\*** | 0.174 | -0.155 | **-0.345\*** | 0.136 |
| 7 Days Ago | CO | **0.448\*** | **0.515\*** | 0.125 | **0.313\*** | **0.346\*** | 0.083 |
|  | NO2 | 0.227 | **0.378\*** | 0.168 | 0.162 | **0.245\*\*** | 0.115 |
|  | PM 10 | **0.293\*\*** | **0.517\*** | **0.292\*\*** | **0.191\*\*\*** | **0.345\*** | **0.196\*\*\*** |
|  | PM 2.5 | **0.261\*\*\*** | **0.399\*** | **0.267\*\*\*** | **0.171\*\*\*** | **0.240\*\*** | **0.196\*\*\*** |
|  | SO2 | -0.207 | -0.004 | **0.492\*** | -0.123 | -0.003 | **0.362\*** |
| 14 Days Ago | CO | 0.066 | -0.051 | -0.177 | 0.058 | -0.018 | -0.117 |
|  | NO2 | -0.165 | **-0.257\*\*\*** | **-0.311\*\*** | -0.106 | **-0.172\*\*\*** | **-0.222\*\*** |
|  | PM 10 | -0.009 | -0.045 | -0.157 | 0.010 | -0.004 | -0.108 |
|  | PM 2.5 | -0.107 | -0.126 | -0.157 | -0.070 | -0.079 | -0.102 |
|  | SO2 | **-0.495\*** | **-0.305\*\*** | -0.220 | **-0.312\*** | **-0.203\*\*** | -0.172 |

\*\*\*, \*\*, \* stands for 10%, 5% and 1% level of significance.

Table 4 highlights the Kendall and Spearman’s correlation coefficients corresponding to 4 weather indicators evaluated at three time frames i.e. same day, 7 days ago and 14 days ago among the daily confirmed, recovered, and deceased cases. The average and maximum temperature at all the three time frames possess strong positive correlation with both daily confirmed and the recovered cases as posited by [32]. The temperatures same day are also significant for the daily deceased cases. The study also highlights the association of average and maximum humidity at two time frames (same day and 7 days ago) with daily deceased cases. The correlation is negative, which implies that as the humidity increases, the daily mortality lowers. The average and maximum humidity at 7 days is also significant for daily recovered cases. By and large, the maximum relative Humidity on the same day doesn’t exhibit a significant association with the COVID-19 daily confirmed cases in the duration of this study, which contradicts the study in Iran [28]. Both the Spearman as well as Kendall correlation coefficients supports the above findings however, the magnitude of correlation coefficients are higher in the Spearman correlation test.

**Table 4:** Correlation coefficients between weather indicators and COVID-19 cases

| Time Frame | Weather Indicators | Spearman Correlation Coefficient |  |  | Kendall Correlation Coefficient |  |  |
| --- | --- | --- | --- | --- | --- | --- | --- |
|  |  | Confirmed Cases | Recovered Cases | Deceased | Confirmed Cases | Recovered Cases | Deceased |
| Same Day | Humidity Maximum | -0.148 | -0.118 | **-0.484\*** | -0.096 | -0.073 | **-0.366\*** |
|  | Humidity Average | 0.017 | -0.039 | **-0.442\*** | 0.029 | -0.013 | **-0.331\*** |
|  | Temperature Maximum | **0.299\*\*** | **0.276\*\*\*** | **0.492\*** | **0.198\*\*\*** | **0.181\*\*\*** | **0.375\*** |
|  | Temperature Average | **0.403\*** | **0.381\*** | **0.473\*** | **0.278\*** | **0.243\*\*** | **0.347\*** |
| 7 Days Ago | Humidity Maximum | -0.093 | **-0.406\*** | **-0.404\*** | -0.062 | **-0.269** | **-0.293\*** |
|  | Humidity Average | 0.084 | **-0.288\*\*** | **-0.391\*** | 0.059 | **-0.185\*\*\*** | **-0.281\*** |
|  | Temperature Maximum | **0.438\*** | **0.672\*** | 0.186 | **0.317\*** | **0.511\*** | 0.126 |
|  | Temperature Average | **0.526\*** | **0.698\*** | 0.187 | **0.365\*** | **0.531\*** | 0.128 |
| 14 Days Ago | Humidity Maximum | -0.193 | **-0.250\*\*\*** | 0.000 | -0.131 | **-0.170\*\*\*** | -0.004 |
|  | Humidity Average | 0.021 | -0.143 | 0.131 | 0.020 | -0.076 | 0.090 |
|  | Temperature Maximum | **0.455\*** | **0.456\*** | -0.024 | **0.302\*** | **0.332\*** | -0.024 |
|  | Temperature Average | **0.536\*** | **0.527\*** | 0.058 | **0.360\*** | **0.396\*** | 0.028 |

\*\*\*, \*\*, \* stands for 10%, 5% and 1% level of significance.

This study reveal a significant correlation between the transmission of COVID-19 outbreaks and the atmospheric pollutants with a combination of specific climatic conditions. This study has shown evidence of pollution and weather indicators correlation with COVID-19 cases; however, there are various limitations under which this study has been conducted. The variables such as lockdown measures, people’s individual immunity, migration index, and other climate indicators can impact the results presented in this study.

## 4. Conclusions

The environment pollution as well as weather indicators can play a crucial role in the fight against coronavirus. To the best of our knowledge, this is the first study to investigate the impact of environment pollution and weather indicators on COVID-19 incidences in Indian context. Unlike other studies, the present study investigates the impact of these indicators in three time frames of same day, 7 days ago and 14 days ago. We have analysed that the pollution indicators are significantly correlated with COVID-19 cases, hence limited human exposure to outdoor pollution may lead to decline in COVID-19 cases. The present study can be further enhanced by including other parameters such as demographic variations, healthcare infrastructure, and social policies like lockdowns to provide better insight into the fight against COVID-19.

## Data Availability

The data that support the findings of this study are available from OpenAQ platform and Weather Underground. These data were derived from the following resources available in the public domain: https://www.covid19india.org/, https://openaq.org and https://www.wunderground.com/. These data sources are referred in the manuscript.

https://openaq.org

https://www.wunderground.com/

## Acknowledgments

This research was supported partially by the research grant scheme of TEQIP-III, MNIT Jaipur.

## Reference

[1] WHO, “Coronavirus disease 2019 (COVID-19) Situation Report – 121,” World Health Organization, 2020.

[2] S. Mahato, S. Pal and K. G. Ghosh, “Effect of lockdown amid COVID-19 pandemic on air quality of the megacity Delhi, India,” Science of The Total Environment, vol. 730, 2020.

[3] WHO, “WHO Global Urban Ambient Air Pollution Database (Update 2016),” 2016.

[4] J. Ciencewicki and I. Jaspers, “Air pollution and respiratory viral infection,” Inhalation Toxicology, vol. 19, pp. 1135–1146, 2007.

[5] J. L. Domingo and J. Rovira, “Effects of air pollutants on the transmission and severity of respiratory viral infections,” Environmental Research, vol. 187, 2020.

[6] P.-S. Chen, F. Tsai, C. Lin, C.-Y. Yang, C.-C. Chan, C.-Y. Young and C.-H. Lee, “Ambient influenza and avian influenza virus during dust storm days and background days,” Environmental Health Perspectives, vol. 118, no. 9, p. 2010, 1211–1216.

[7] R. Glass and J. Rosenthal, “International approach to environmental and lung health a perspective from the fogarty international center,” Annals of the American Thoracic Society, vol. 15, pp. S109–S113, 2018.

[8] Y. Cui, Z.-F. Zhang, J. Froines, J. Zhao, H. Wang, S.-Z. Yu and R. Detels, “Air pollution and case fatality of SARS in the People’s Republic of China: an ecologic study,” Environmental Health, vol. 2, 2003.

[9] L. Martelletti and P. Martelletti, “Air Pollution and the Novel Covid-19 Disease: a Putative Disease Risk Factor,” Clinical Medicine, vol. 15, pp. 1–5, 2020.

[10] E. Conticini, B. Frediani and D. Caro, “Can atmospheric pollution be considered a cofactor in extremely high level of SARS-CoV-2 lethality in Northern Italy?,” Environmental Pollution, vol. 261, 2020.

[11] Y. Zhu, J. Xie, F. Huang and L. Cao, “Association between short-term exposure to air pollution and COVID-19 infection: Evidence from China,” Science of The Total Environment, vol. 727, 2020.

[12] Y. Ogen, “Assessing nitrogen dioxide (NO2) levels as a contributing factor to coronavirus (COVID-19) fatality,” Science of The Total Environment, vol. 726, 2020.

[13] J. Bontempi, “First data analysis about possible COVID-19 virus airborne diffusion due to air particulate matter (PM): the case of Lombardy (Italy),” Environmental Research, vol. 186, 2020.

[14] X. Wu, R. Nethery, B. Sabath, D. Braun and F. Dominici, “Exposure to Air Pollution and COVID-19 Mortality in the United States,” medRxiv, 2020.

[15] Y. Yao, J. Pan, W. Wang, Z. Liu, H. Kan, X. Meng and W. Wang, “Spatial Correlation of Particulate Matter Pollution and Death Rate of COVID-19,” medRxiv, 2020.

[16] D. Fattorini and F. Regoli, “Role of the atmospheric pollution in the Covid-19 outbreak risk in Italy,” Environmental Pollution, vol. 264, 2020.

[17] M. F. Bashir, B. J. MA, Bilal, B. Komal, M. A. Bashir, T. H. Farooq, N. Iqbal and M. Bashir, “Correlation between environmental pollution indicators and COVID-19 pandemic: A brief study in Californian context,” Environmental Research, vol. 187, 2020.

[18] J. Yuan, H. Yun, W. Lan, W. Wang, S. G. Sullivan, S. Jia and A. H. Bittles, “A climatologic investigation of the SARS-CoV outbreak in Beijing, China,” American Journal of Infection Control, vol. 34, no. 4, pp. 234–236, 2006.

[19] J. Shaman, E. Goldstein and M. Lipsitch, “Absolute Humidity and Pandemic Versus Epidemic Influenza,” American Journal of Epidemiology, vol. 173, no. 2, p. 127–135, 2011.

[20] M. F. Bashir, B. Ma, B. B. Komal, M. A. Bashir, D. Tan and M. Bashir, “Correlation between climate indicators and COVID-19 pandemic in New York, USA,” Science of The Total Environment, vol. 728, 2020.

[21] Á. Briz-Redón and Á. Serrano-Aroca, “A spatio-temporal analysis for exploring the effect of temperature on COVID-19 early evolution in Spain,” Science of The Total Environment, 2020.

[22] H. Qi, S. Xiao, R. Shi, M. P. Ward, Y. Chen, W. Tu, Q. Su, W. Wang, X. Wang and Z. Zhang, “COVID-19 transmission in Mainland China is associated with temperature and humidity: A time-series analysis,” Science of The Total Environment, 2020.

[23] S. Gupta, G. S. Raghuwanshi and A. Chand, “Effect of weather on COVID-19 spread in the US: A prediction model for India in 2020,” Science of The Total Environment, vol. 728, 2020.

[24] Y. Ma, Y. Zhao, J. Liu, X. He, B. Wang, S. Fu, J. Yan, J. Niu, J. Zhou and B. Luo, “Effects of temperature variation and humidity on the death of COVID-19 in Wuhan, China,” Science of The Total Environment, vol. 724, 2020.

[25] J. Liu, J. Zhou, J. Yao, X. Zhang, L. Li, X. Xu, X. He, B. Wang, S. Fu, T. Niu, J. Yan, Y. Shi, X. Ren, J. Niu, W. Zhu, S. Li, B. Luo and K. Zhang, “Impact of meteorological factors on the COVID-19 transmission: A multi-city study in China,” Science of The Total Environment, vol. 726, 2020.

[26] P. Shi, Y. Dong, H. Yan, C. Zhao, X. Li, W. Liu, M. He, S. Tang and S. Xi, “Impact of temperature on the dynamics of the COVID-19 outbreak in China,” Science of The Total Environment, 2020.

[27] M. Şahin, “Impact of weather on COVID-19 pandemic in Turkey,” Science of the Total Environment, vol. 728, 2020.

[28] M. Ahmadi, A. Sharifi, S. Dorosti, S. J. Ghoushchi and N. Ghanbari, “Investigation of effective climatology parameters on COVID-19 outbreak in Iran,” Science of The Total Environment, 2020.

[29] B. Oliveiros, L. Caramelo, N. C. Ferreira and F. Caramelo, “Role of temperature and humidity in the modulation of the doubling time of COVID-19 cases,” medRxiv, 2020.

[30] M. Kendall, “A New Measure of Rank Correlation,” Biometrika, vol. 30, pp. 81–89, 1938.

[31] C. Spearman, “The proof and measurement of association between two things,” The American Journal of Psychology, vol. 15, no. 1, pp. 72–101, 1904.

[32] R. Tosepu, J. Gunawan, D. S. Effendy, L. O. A. I. Ahmad, H. Lestari, H. Bahar and P. Asfian, “Correlation between weather and Covid-19 pandemic in Jakarta, Indonesia,” Science of the Total Environment, vol. 725, 2020.

